# Feasibility of an App-Assisted and Home-Based Video Version of the Timed up and Go Test for Patients with Parkinson Disease: vTUG

**DOI:** 10.1101/2024.09.30.24314647

**Authors:** Marcus Grobe-Einsler, Anna Gerdes, Tim Feige, Vivian Maas, Clare Matthews, Alejandro Mendoza García, Laia Comas Fages, Elin Haf Davies, Thomas Klockether, Björn H. Falkenburger

## Abstract

**Background:** Parkinson Disease (PD) is a progressive neurological disorder. Current therapeutic trials investigate treatments that can potentially modify the disease course. Testing their efficiency requires frequent and precise clinical outcome assessments (COA) of symptoms that remain problematic under symptomatic treatments, such as gait and balance. Home-based examinations may enhance patient compliance and, in addition, produce more reliable results by assessing patients more regularly in their familiar surroundings.

**Objective:** The objective of this study was to assess the feasibility of a digitized COA designed to video record the Timed up and Go (vTUG) test at home via a study-specific smartphone app for patients with PD.

**Method:** In this study, 28 patients were recruited and asked to perform at home each week a set of three consecutive vTUG tests over a period of 12 weeks using an app. The videos were subjected to a manual review to ascertain the durations of the individual vTUG phases, as well as to identify any errors or deviations in the setup that might have influenced the result. To evaluate the usability and user-friendliness of the vTUG and app, the System Usability Scale (SUS) and User Experience Questionnaire (UEQ) were administered to patients at the study end.

**Results:** Overall, 19 patients completed the 12-week study, 17 of which recorded 10 videos or more. A total of 706 vTUGs with complete timings were recorded. Random Forest Regression yielded “time to walk up” as the most important segment of the vTUG for predicting the total time. Variance of vTUG total time was significantly higher between weeks than it was between the three consecutive vTUGs at one time point [F(254,23) = 6.50, p < 0.001]. The correlation between vTUG total time and UPDRS III total score was weak (r = 0.24). Yet, correlation between vTUG and a derived gait subscore (UPDRS III items 9-13) was strong (r = .59). A linear mixed-effects model revealed a significant effect of patient-reported motion status on vTUG total time. Including additional variables such as UPDRS III gait subscore, different footwear, and chairs used, further improved the model fit.

**Conclusion:** Assessment of gait and balance by home-based vTUG is feasible. Factors influencing the read-out were identified and could be controlled for future use and longitudinal trials.

## Introduction

Parkinson disease (PD) has the fastest growing prevalence of all neurologic diseases worldwide^1^. Motor symptoms are a major driver of reduced quality of life and are caused by degeneration of dopaminergic neurons in the substantia nigra. Symptomatic treatment strategies therefore focus on elevation of dopamine levels in the brain. First disease-modifying treatments are under investigation. These therapies aim to delay the disease progression and should therefore be applied in early stages of the disease. Several obstacles for planning such clinical trials remain: Reliable and sensitive outcome measures are required to detect the subtle changes of symptom severity expected in early-stage PD. Frequent travels for repeated neurological examinations are challenging for patients with movement disorders. The resultant exhaustion and even the clinical environment itself influence the results of the assessments and some authors have argued that patients with PD (PwPD) should preferably be tested in their own home environment, whenever possible^2–5^. Utilization of Digital Health Technologies (DHT) may help overcome these obstacles. They are potentially more sensitive to subtle changes of motor function compared to conventional clinical scales^6^ and can be applied in a remote setting for longitudinal monitoring. The benefits of DHTs in monitoring PwPD are increasingly recognized in the field, with the coronavirus pandemic acting as an additional catalyst for their application as remote assessments^7,8^.

To optimize patient adherence and completeness of data, a remote performance outcome measure (PerfO) should be well accepted in the field and easy to apply. The “Timed Up and Go”-Test (TUG) qualifies for this purpose^3,9^. In this test, the patient is asked to stand up from a chair, walk three meters in a straight line, turn around, walk back, and sit back down. The objective assessment is made by measuring the time to complete the task. It was originally designed to evaluate dynamic balance, functional mobility and risk of falls in geriatric patients^10,11^, but was later demonstrated to be a valuable and efficient method for evaluation of mobility in PwPD^12,13^ with a high test-retest reliability^14,15^. Several approaches to digitize the TUG with sensors have been introduced, which are summarized under the term instrumented TUG (iTUG)^16^. The sensors allow for accurate subdivision of the TUG phases by postural transitions, and introduce qualitative gait analysis, which ultimately leads to improved discriminatory properties in early disease stages^16,17^. However, the requirement for additional hardware limits the feasibility and increases burden for patients. Additionally, unsupervised digital assessments at home using sensors leave the investigator blind for unsuspected sources of variability. For example, the walking speed may be biased by the size of a room and furniture, while the time required to rise from a chair depends on the size and type of chair^18^. These factors can easily be identified by video based PerfOs. Considering the current advances in camera technology, a TUG assessment using a smartphone seems promising. The objective of this study was to develop and validate a self-applied, video recorded, assisted remote TUG for home application, named vTUG, via a smartphone app.

## Methods

### Recruitment

In total, 33 PwPD were recruited in the German Center for Neurodegenerative Diseases at the Bonn and Dresden sites between January and December 2023. Five patients participated in the in-clinic feasibility assessment; 28 patients participated in the home-based longitudinal part. Inclusion criteria were i) individuals aged 18 and above who met the diagnostic criteria for PD as stipulated by the Movement Disorder Society^19^; ii) Hoehn and Yahr Stages 1 to 4 and iii) the ownership of an Android or iOS Smartphone with internet access and iv) ability to comply with study protocol without the risk of falling according to the investigator. Exclusion criteria were severe comorbidities that could interfere with assessments (for example dementia, risk of falling or severe psychiatric disease).

### Ethics Approval

This study was conducted in accordance with the Declaration of Helsinki. The study was approved by the local ethics committees (BO-EK-149032021_3) and written informed consent was obtained from all patients before the participation.

### App Implementation

The vTUG module was implemented in an e-health app (Atom5™ by Aparito), with compatibility for Android and iOSv to enable patients to use their own phone or tablet. After initialisation via a unique patient identifier QR code, the activated module contained instructional videos and text on how to perform the vTUG, available in both English and German. The actual performance was recorded within the app and uploaded for central review and assessment. Before completing the vTUG assessment, patients were asked to respond to questionnaires from two categories: The first was a „health thermometer“ (numeric value from 0-100) as a measure for general health. The second consisted of a self-evaluation of motion (ON or OFF), the time since last dopaminergic medication and changes in medication since the last assessment. At the end of the study, patients were asked to respond to the User Experience Questionnaire (UEQ)^20^ and the System Usability Scale (SUS)^21^. The SUS is a widely used Likert-Scale and ten-item questionnaire with five possible response options. The SUS is more focused on evaluating the usability of a system, while the UEQ has a broader scope, encompassing the overall user experience. The UEQ assesses Attractiveness, Perspicuity, Efficiency, Dependability, Stimulation, and Novelty of a technical system. Patients received push messages via the app as reminders for upcoming assessments for the duration of the study.

### Study Design and Setup

Clinical information was collected during the baseline visit in the study center and included age, sex, Hoehn and Yahr stages, and disease onset. The MDS-UPDRS III scores were obtained in Bonn from a longitudinal cohort study in which the participants took part, and in Dresden from the medical records of the Department of Neurology at the University Hospital. Each score corresponds to the day of recruitment. A *UPDRS III gait subscore* was calculated as the sum of items 9-13 of the MDS-UPDRS III.

The first stage of the project was an in-clinic usability study to assess if patients were able to follow the in-app instructions provided to perform the vTUG task independently. Five patients were asked to navigate the app by themselves, under the surveillance of a clinician. The clinician noted difficulties that the patients experienced and whether they required help. Feedback regarding user friendliness from this phase was subsequently implemented in the app. This included bigger font size, easier navigation through the app, optimization of the instructions and translations. The second stage consisted of a longitudinal home monitoring. The investigations proceeded as follows: The app installation and initialisation process and first assessment were carried out at the study site under the supervision of an investigator, who provided advice if requested. To start the assessment, patients were asked to watch the video instructions, respond to the first questionnaires and prepare the set-up as follows: A floor-mark was placed at 3 m distance from a chair. A provided tripod with the mounted smartphone was placed another 2 m along the same line. The chair, floor mark and tripod were to be aligned without any obstacles in between. The height and position of the smartphone was adjusted to capture the standing patient on the 3 m floor mark from „head-to-heel“. The patient was then asked to sit down in the chair to start the assessment. The recording was initiated either beforehand by the patient, via voice command when the patient is already sitting in the chair, or via a second person. During the vTUG assessments, patients stood up (without assistance of the arms, if possible), walked 3 m at normal walking speed, turned around, walked back, and sat back down again. This sequence was repeated three times and each recorded on video. While the first session was performed under supervision of an investigator, the consecutive weekly questionnaires and recordings were performed independently by patients at home, once a week for 12 weeks, again in triplicates. To avoid consecutive erroneous performances, the first home recording was centrally reviewed by an investigator in the same week and patients were contacted to correct performance in the following recording, if necessary. Additionally, patients were able to contact investigators from their local study site if they had questions.

### Data Analysis

All videos underwent manual quality control for completeness of the recording and correct framing of the person. The five stages making up the TUG (stand, walk up, turn 180°, walk back and turn to sit) were timed using definitions developed to identify the start and end of each stage. The duration of pauses between stages and the total time taken were also recorded. For each patient it was also noted how many times and in which videos they were in a different location, wearing different footwear or using a different chair from the original setup.

Participants performed the vTUG three times consecutively at each time point, resulting in three individual vTUG measurements per session. The mean of these three trials was calculated to represent the participant’s performance at that time point.

#### Relevance of vTUG-stages on total time

To assess the relevance of the five TUG stages described above on the total TUG time, we calculated both Pearson correlation coefficients and performed a Random Forest regression analysis. For the Random Forest, we conducted a grid search to identify the optimal hyperparameters for the minimum number of samples per leaf and the number of trees in the forest.

#### Correlation with UPDRS III

To evaluate the suitability of the home-based vTUG for measuring motion and particularly gait impairments in people with Parkinson’s disease, we calculated Pearson correlation coefficients and performed individual linear regressions of vTUG total time on the UPDRS III total score and UPDRS III gait score.

#### Variance

To assess the vTUG’s ability to detect changes in walking performance, we calculated the within-session variance among the three vTUG trials conducted at each time point, as well as the variance between these mean values over the 12-week study period. To evaluate whether the variances differed significantly within and between time points, we performed an F-test, calculating the F-statistic and corresponding p-value.

#### Influences on total time

To identify relevant variables for predicting the dependent variable *vTUG total time* we estimated a mixed linear model using the *mixedlm()* function from the *statsmodels* package (v. 0.14.2) in Python with the *UPDRSIII total*, *UPDRSIII gait subscore*, patients’ *perceived motor status*, *age*, *disease duration* as well as *shoes worn, chairs used* and *locations filmed at* as predictor variables. We included random intercepts for each participant to account for the repeated measures design, allowing us to control for individual variability in baseline performance. We employed a stepwise model-building procedure, sequentially adding the predictor variables in the order listed above. At each step, we compared the Akaike Information Criterion (AIC) and Bayesian Information Criterion (BIC) and conducted a Likelihood Ratio Test (LRT) between the reduced model (excluding the new variable) and the full model (including the new variable) to determine whether to retain the variable in the model. In order to calculate AIC, BIC and perform LRTs we estimated all models using maximum likelihood.

To assess the final model fit, we calculated the marginal and conditional R² values using Nakagawa and Schielzeth’s formula^22^.

#### Usability

To assess the usability of the vTUG we applied the System Usability Score (SUS)^21^ and the User Experience Questionnaire (UEQ).

The SUS provides a subjective assessment of usability from the patient’s perspective through ten Likert scale questions. For odd-numbered questions, participants rate from 1 (strongly disagree) to 5 (strongly agree), while even-numbered questions are rated inversely from 5 to 1. The SUS score is calculated by adjusting the responses—subtracting 1 from each odd-numbered question and subtracting the response from 5 for each even-numbered question. These adjusted scores are summed and multiplied by 2.5, resulting in a total score ranging from 0 to 100. Higher scores indicate better perceived usability, with scores above 68 considered above average and those exceeding 80 considered excellent.^23^

The UEQ is a standardized questionnaire designed to assess the user experience of products, systems, or services. It consists of 26 items that measure six key dimensions: Attractiveness, Perspicuity (clarity), Efficiency, Dependability, Stimulation, and Novelty. Participants rate each item on a seven-point scale ranging from −3 (extremely bad) to +3 (extremely good). Instead of generating an overall score, mean values for each domain are analyzed, with values greater than 0.8 considered a positive evaluation.^24^

## Results

In total, 28 patients were included in this study, with three patients dropping out after the in-clinic assessment due to personal reasons that were not linked to the burden of the study. The clinical and demographic data for the remaining 25 patients are summarized in Table 1.

**Table 1.** Baseline Characteristics.

| **Baseline characteristic** |  |  |
| --- | --- | --- |
| Total Participants | 25 |  |
|  | *Mean* | *SD* |
| MDS UPDRSIII total | 20.5 | 12.2 |
| MDS UPDRSIII gait subscore* | 2.6 | 2.6 |
| Number of Weeks Recorded | 9.8 | 2.4 |
| vTUG Rounds per Patient | 26.3 | 7.5 |
| vTUG Triplets per Patient | 7.5 | 2.7 |
|  | *Median* | *[Q1-Q3]* |
| age | 61.0 | [58.0-66.0] |
| age at disease onset | 55.0 | [51.0-59.0] |
|  | *Count* | *Percent* |
| sex |  |  |
| f | 8 | 32 |
| m | 17 | 68 |
| Hoehn & Yahr |  |  |
| 1 | 6 | 24 |
| 2 | 17 | 68 |
| 3 | 2 | 8 |

\* We calculated a gait subscore as a sum of MDS UPDRS III Items 9-13

### Video summary

A total of 273 videos were recorded, of which 262 passed quality control and were included in the analysis. Four videos were unavailable due to upload failures, and seven were excluded because the camera was partially covered or did not capture a vTUG at all.

Eight patients completed the task every week, submitting 12 videos each that included 36 vTUGs. Eleven additional patients also completed the 12-week period but missed some weekly video recordings due to unspecified issues.

Over the course of the study, we observed only a small attrition rate, with the percentage of available data decreasing from 88% in week 1 to 76% in week 12. Linear regression analysis estimated a 1.3% decrease in available data per week.

The 262 videos contained 784 vTUG assessments (two videos contained only two instead of the standard three vTUGs). Out of these, 706 covered the entire task; timing was incomplete in 78 vTUGs because the recordings failed to capture the beginning of the first TUG test.

In nine videos, the participants’ feet were out of frame at the 3-meter mark, mostly due to walking past the floor mark or improper setup. In 12 videos—seven of which were recorded by a single patient—the participants’ shoulders were out of frame during the turn at the 3-meter mark.

Most patients consistently used the same chair at home, with only a few changing it once or twice. The most frequent changes were observed in footwear: only nine patients consistently used the same pair of shoes, while the others changed their shoes up to seven times. Additionally, the majority of patients recorded each TUG at the same location, with eight patients changing locations once or twice. The variability in vTUG total times based on different shoes, chairs, and locations used by the participants is visualized in Figure 2.

**Figure 1:**
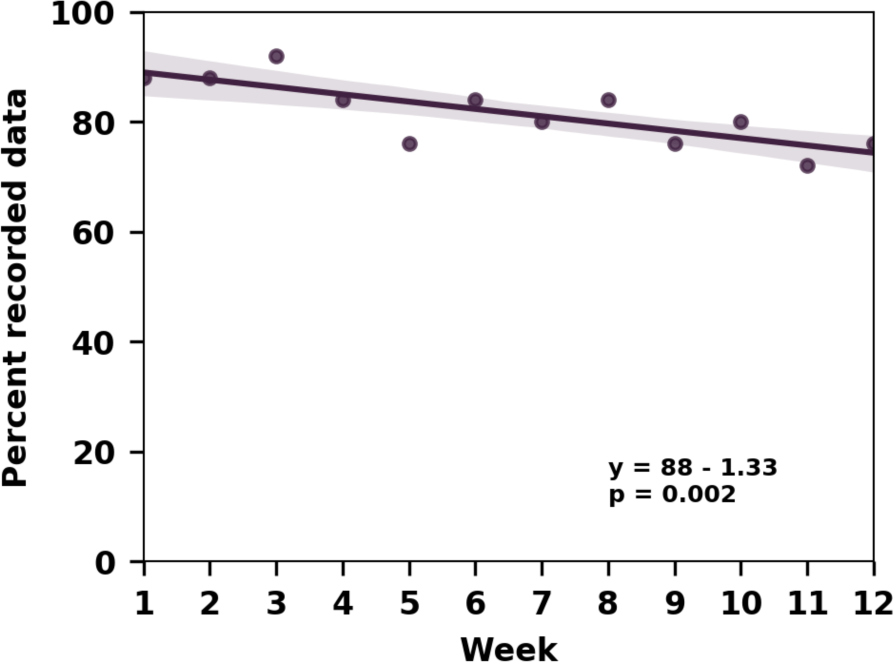
Despite a small attrition rate, data availability remained quite high throughout the study duration.

**Figure 2.**
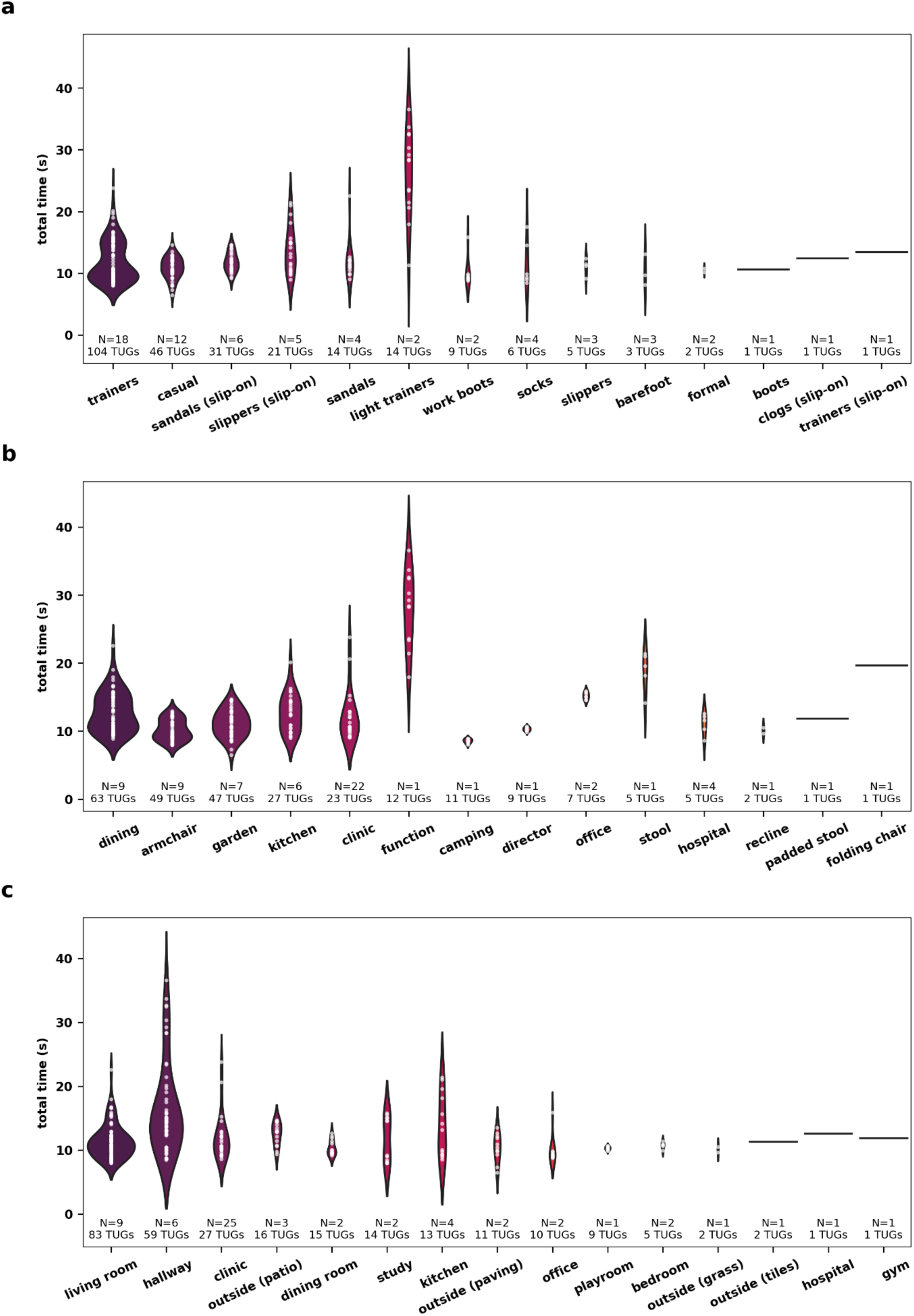
Different shoes, chairs, and locations introduce notable variability in vTUG total time. Violin plots display the distribution of vTUG total time by the shoes worn (a), chairs used (b), and locations filmed at (c). The number of patients per category (N) and the total number of TUGs performed are indicated for each category.

### vTUG Timing Data

The average time required to complete a vTUG was 12.5 (4.7) seconds. Figure 3 illustrates the range of total times across all patients and the number of vTUGs recorded. The patient with the longest average time during the study period took 27.6 seconds on average and also recorded the longest individual vTUG time of 38.7 seconds. In contrast, the patient with the fastest average time completed the vTUG in 8.6 seconds, with the single fastest time being 6.2 seconds. Table 2 provides an overview of the timings for each individual vTUG segment.

**FIGURE 3:**
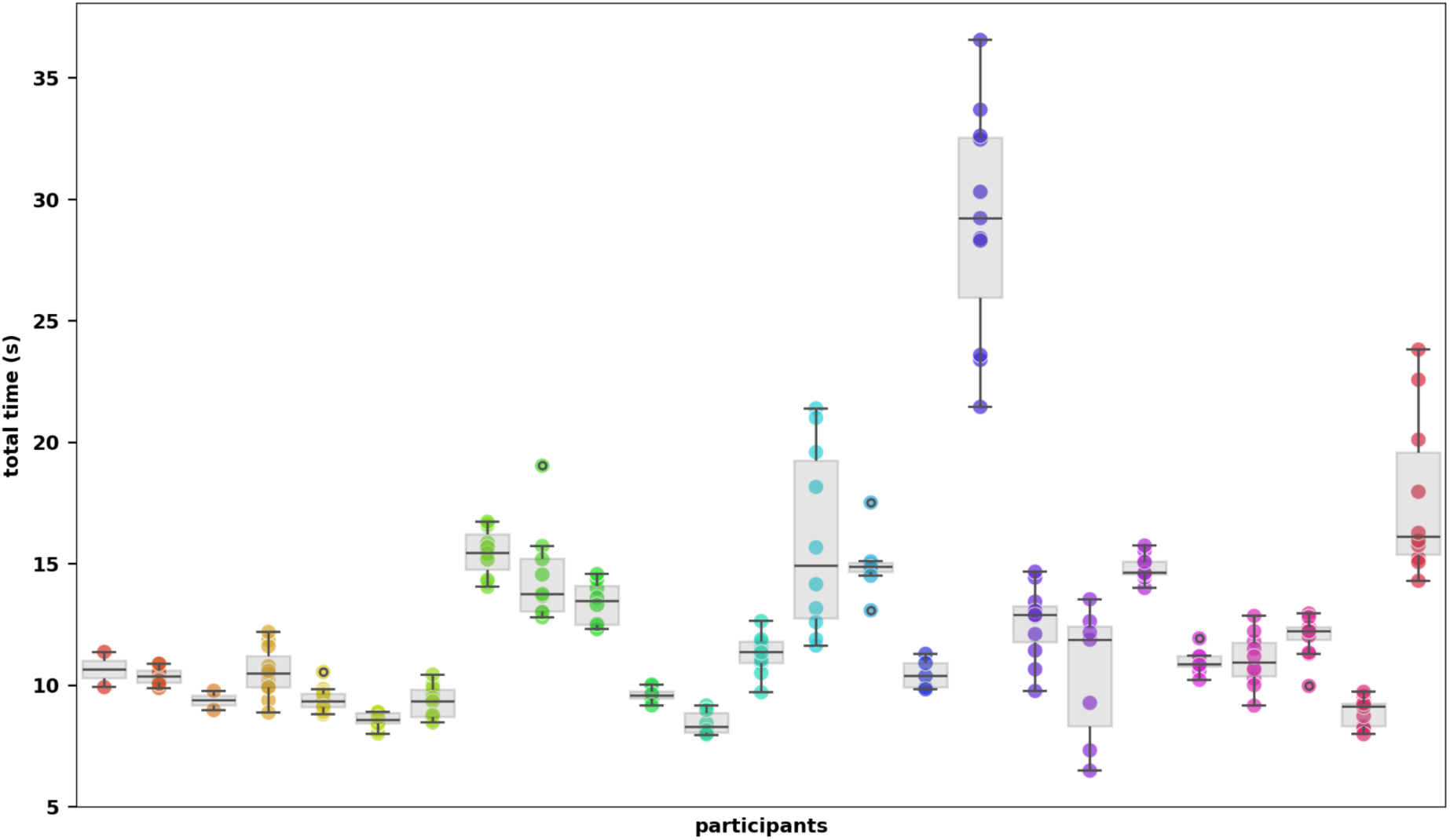
vTUG total time displayed for all participants. *Different colours represent individual participants*.

**Table 2.** descriptive Statistics for recorded vTUG segments.

| vTUG segment | N | Mean (SD) | Min | Max |
| --- | --- | --- | --- | --- |
| time to stand | 706 | 1.29 (0.66) | 0.57 | 8.38 |
| time to walk up | 742 | 3.43 (1.17) | 1.63 | 10.21 |
| time to turn 180 | 757 | 2.1 (0.82) | 1.03 | 7.4 |
| time to walk back | 757 | 2.68 (0.91) | 0.87 | 5.92 |
| time to turn & sit | 757 | 2.81 (1.70) | 1.33 | 15.69 |
| total time | 706 | 12.51 (4.72) | 6.17 | 38.67 |

all time measures are in seconds

**Table 3.** Linear mixed effect models evaluating the influence of clinical and environment variables on vTUG total time.

| Fixed effects | Estimates (95 CI) | Std. Error | p |
| --- | --- | --- | --- |
| Intercept | 11.507 (9.921 - 13.093) | 0.809 | < 0.001 |
| patient-reported motion status<br>(reference category: on) | 1.216 (0.487 - 1.944) | 0.372 | 0.001 |
| UPDRSIII gait | 0.381 (-0.018 - 0.780) | 0.204 | 0.061 |
| Shoe Type<br>(reference category: trainers) |  |  |  |
| barefoot | -1.173 (-3.271 - 0.925) | 1.07 | 0.273 |
| casual | -0.440 (-1.668 - 0.788) | 0.626 | 0.482 |
| clogs (slip-on) | -0.114 (-3.654 - 3.425) | 1.806 | 0.949 |
| formal | -0.157 (-2.601 - 2.287) | 1.247 | 0.9 |
| light trainers | 0.077 (-3.747 - 3.900) | 1.951 | 0.969 |
| sandals | 0.900 (-0.730 - 2.529) | 0.831 | 0.279 |
| sandals (slip-on) | 0.740 (-0.698 - 2.177) | 0.733 | 0.313 |
| slippers | 0.441 (-1.720 - 2.601) | 1.102 | 0.689 |
| slippers (slip-on) | 0.692 (-1.047 - 2.432) | 0.888 | 0.435 |
| socks | 0.573 (-1.261 - 2.408) | 0.936 | 0.54 |
| (trainers (slip-on) | 1.755 (-1.596 - 5.106) | 1.71 | 0.305 |
| work boots | 0.197 (-2.097 - 2.491) | 1.17 | 0.866 |
| Chair Type<br>(reference category: dining chair) |  |  |  |
| armchair | -2.235 (-3.950 - -0.520) | 0.875 | 0.011 |
| camping | -2.600 (-5.455 - 0.255) | 1.457 | 0.074 |
| clinic | 2.073 (-0.526 - 4.672) | 1.326 | 0.118 |
| director | -2.576 (-5.078 - -0.073) | 1.277 | 0.044 |
| function | 12.092 (5.922 - 18.263) | 3.148 | 0.0 |
| garden | -1.406 (-3.143 - 0.332) | 0.887 | 0.113 |
| hospital | -1.455 (-4.375 - 1.466) | 1.49 | 0.329 |
| kitchen | -1.460 (-2.794 - -0.127) | 0.68 | 0.032 |
| office | 0.349 (-2.375 - 3.073) | 1.39 | 0.802 |
| padded stool | -0.984 (-4.860 - 2.892) | 1.978 | 0.619 |
| recline | -1.246 (-4.442 - 1.951) | 1.631 | 0.445 |
| stool | 4.565 (2.208 - 6.923) | 1.203 |  |

Data are unstandardized coefficients and (95% confidence intervals)

#### Relevance of vTUG-stages on total time

The time to walk up showed the strongest correlation with the total time (r = 0.91), while the times to turn 180 degrees and to turn and sit were also strongly correlated (both at r = 0.89). These three segments of the vTUG were also identified as the most important predictors in the random forest regression.

We performed an 80/20 train-test split to assess the accuracy of the trained model. A grid search for optimal hyperparameters suggested setting the minimum samples per leaf to 1 and using 100 trees in the forest. With these hyperparameters, the random forest was able to predict the vTUG total time with a mean absolute error (MAE) of 0.31 and a mean squared error (MSE) of 0.36. Figure 4 displays both the feature importance plot and the Pearson correlation matrix.

**Figure 4:**
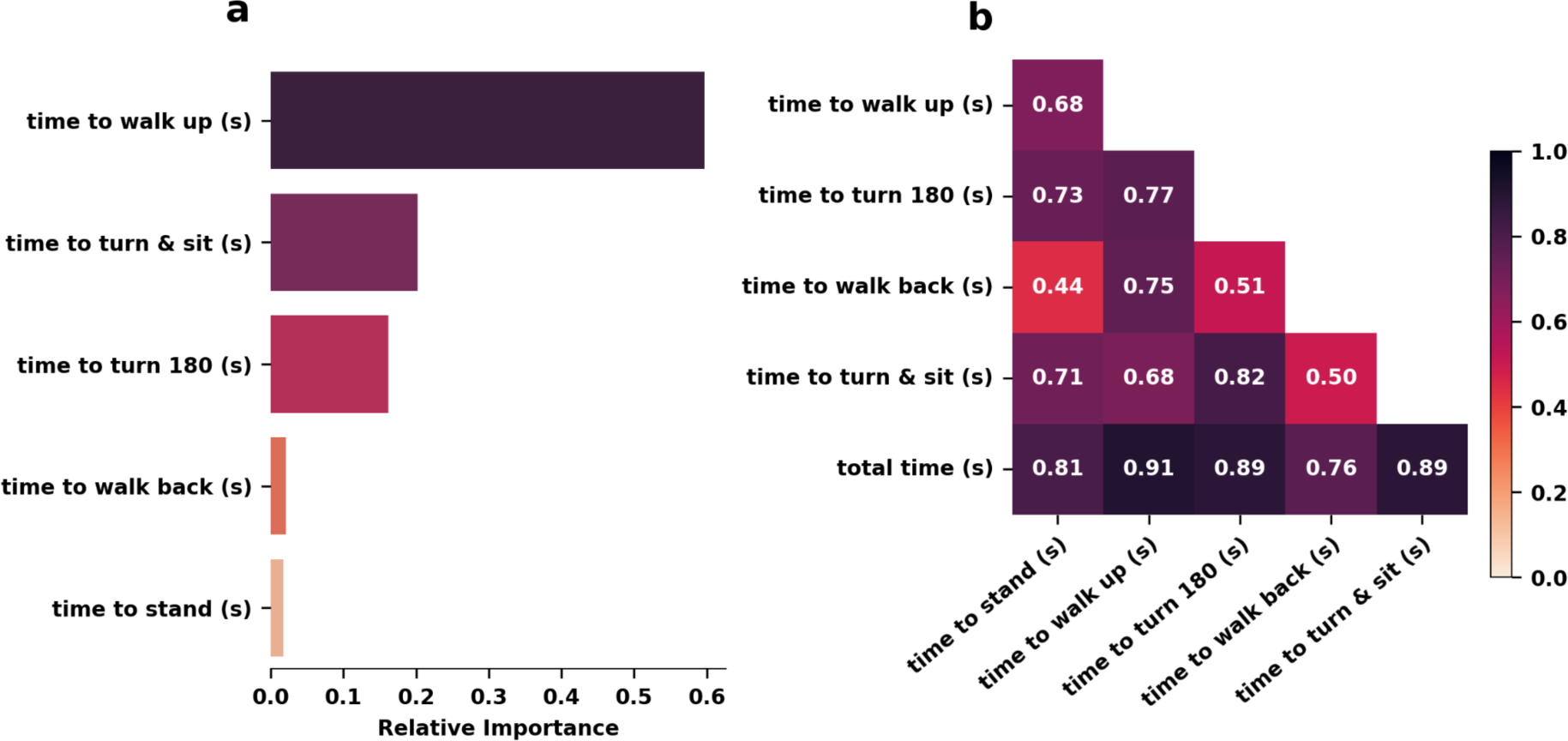
Time to walk up has the highest impact on vTUG total time. a. Relative importance of vTUG segments for predicting vTUG total time. b. Pearson correlations of vTUG segments and vTUG total time.

**Figure 5.**
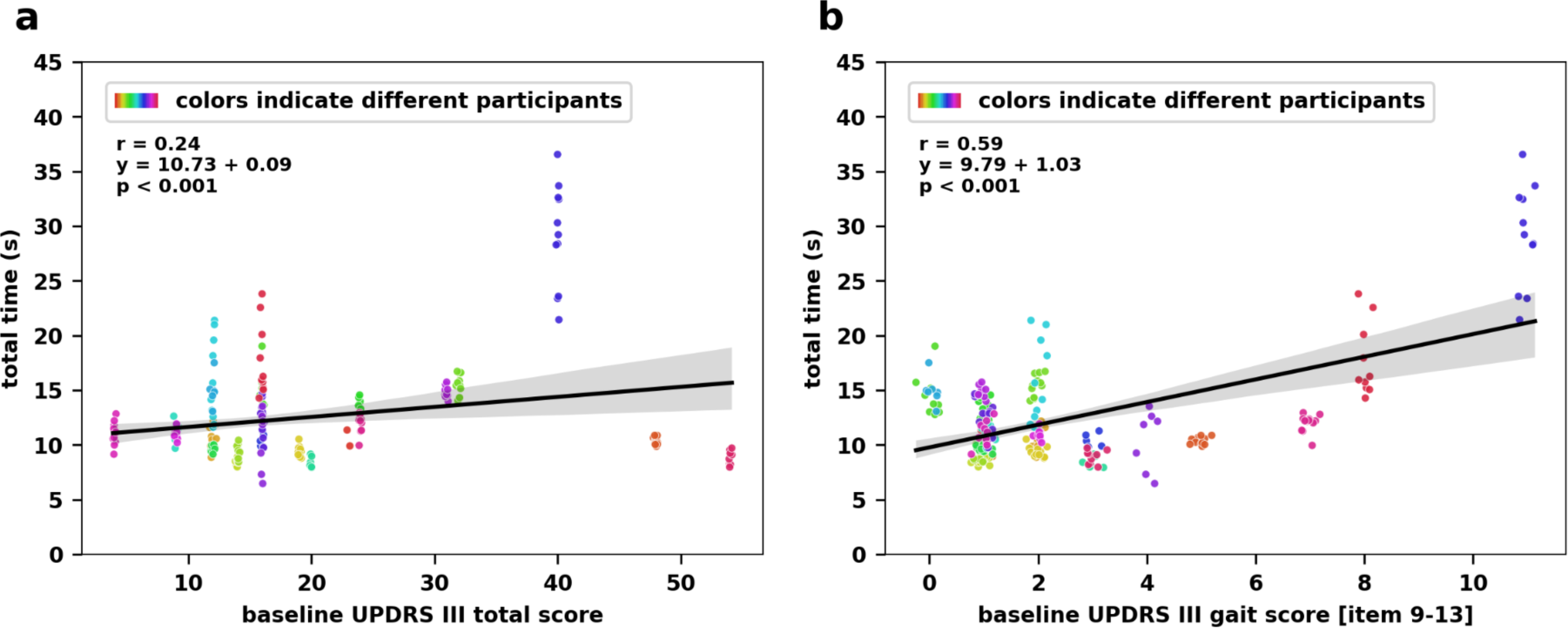
Strong correlation with MDS UPDRS III gait score shows vTUG’s ability to detect gait impairments in people with PD. Regression plots for vTUG total time against baseline MDS UPDRS III total score (a) and gait subscore derived from the sum of UPDRS III items 9-13 (b).

**Figure 6:**
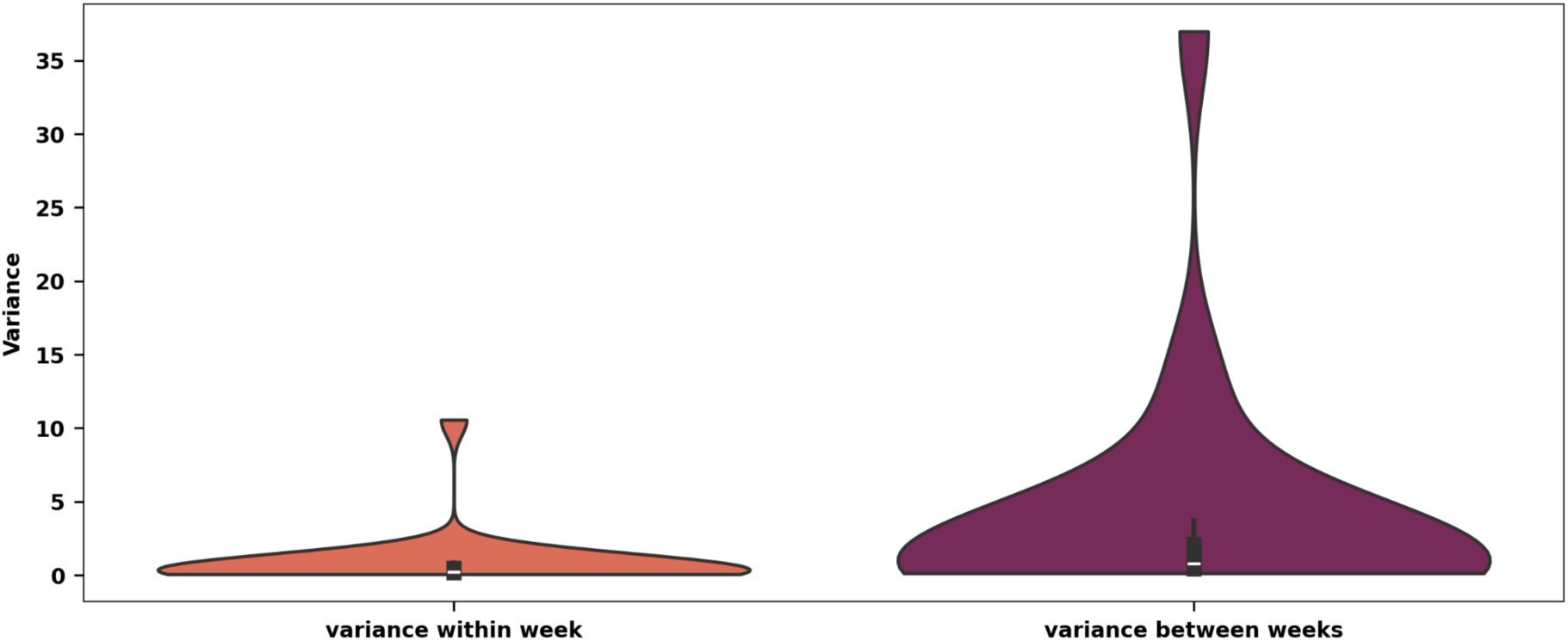
Significant variability emerges in vTUG performance over time, with between-week variance far exceeding within-week consistency. *Variances within a single week for three consecutive vTUG tests and variances between the weekly means of vTUG tests over the 12-week study period are depicted in s^2^*.

#### Relationship between vTUG and MDS UPDRS III

The *vTUG total time* only had a weak correlation with the *MDS UPDRS III total score* (r = 0.24). However, correlation with the derived *MDS UPDRS III gait subscore* was strong (r = 0.59). Linear regression yielded the following equation: y = 10.83 + 0.09x for the *MDS UPDRS III total score* and y = 9.79 + 1.03 for the *MDS UPDRS III gait subscore*.

#### Variance

The variance of the total time in seconds within the three vTUGs performed consecutively in one week had a mean of 0.87 seconds² (SD = 3.19), ranging from 0.01 to 38.6 seconds². In contrast, the variance of the weekly vTUG means over the course of the 12 weeks of the study had a mean of 12.55 seconds² (SD = 4.66), ranging from 6.47 to 36.55 seconds². To assess whether there were significant differences between these variances, an F-test was conducted. The test yielded an F-statistic of 6.50 (p<0.001) indicating a significant difference in vTUG variance within one measuring time point and between weeks.

Recognizing that variance is highly sensitive to outliers, we performed a secondary analysis by excluding vTUGs with total times exceeding 20 seconds, which accounted for only 20 vTUGs. In this filtered dataset, the mean variance within a week was reduced to 0.38 seconds² (SD = 0.61), ranging from 0.01 to 3.81 seconds². The mean variance between weeks was 11.74 seconds² (SD = 2.90). An F-test on this dataset resulted in an F-statistic of 34.61 (p < 0.001), reinforcing the significant difference in variance of the three vTUGs performed within one measurement time point compared to between weeks.

#### Influences on vTUG total time

Among the tested variables, the *UPDRS III gait subscore*, *patient-reported motion status*, *shoe type*, and *chair used* significantly improved the AIC, reducing it from 998.19 in the base model to 938.57 in the full model, and were highly significant in the likelihood ratio test (LRT) (p < 0.05). In contrast, the *UPDRS III total score*, *age*, *disease duration*, *time since last medication*, and *filming location* did not significantly improve the model.

Interestingly, the BIC increased from 1008.43 in the base model to 1036.85 in the full model. This increase may be attributed to BIC’s penalization of models with a high number of parameters, especially those involving categorical variables with multiple levels, such as *shoe type* and *chair used*.

Notably, adding *chair used* to the model led to the *UPDRS III gait subscore* no longer being significant (p = 0.06). Excluding the *UPDRS III gait subscore* from the final model affected the fit indices differently: the AIC decreased slightly to 939.78, while the BIC decreased to 1034.68. The LRT for the full model was just not significant (p = 0.07). Given the mixed changes in the fit indices and the p-value being close to significance, we decided to retain the UPDRS III gait subscore in the full model.

### End-of-study questionnaires

Twenty-one participants completed the end-of-study questionnaire The mean SUS score was 75,5 (SD = 15.04). The UEQ results indicate that perspicuity received the highest mean score of 1.45 (SD=1.14), followed by attractiveness with a mean of 0.9 (SD=0.99). Stimulation had the lowest ranking with a mean score of 0.5 (SD=0.97). Compared to previous studies^25^, perspicuity scored above average, whereas the others scored below average.

## Discussion

### Principal Findings

The study was completed by the majority of patients over the period of three months. Despite some errors, a significant number of evaluable vTUGs were uploaded, demonstrating the system’s usability and patients’ ability to use it, while also generating interpretable data. Furthermore, we identified factors that influence the vTUG performance and hence should be addressed for future work in order to sustain standardization in the home environment.

### Video Data

The measured times demonstrate significant variance from one week to the next. This variance is likely also observed in clinic. Repeated home measurements can potentially compensate for this variance and improve the accuracy of measuring disease progression. In addition, the time taken to complete the vTUG can identify individuals at risk of falling. A threshold of 11.5 s has been suggested^26^. Indeed, the TUG is also a reliable measure in studies of other illnesses than PD, including Lambert-Eaton-Myasthenia^27^ and Essential Tremor^28^. It should be noted that the TUG was initially developed for the purpose of assessing the risk of falls and functional gait disorders in geriatric patients^29^. This also provides insight into an individual’s mobility, which has been proven to have a significant impact on health, including brain health. Consequently, our study may also serve as a foundation for the application of the vTUG in geriatric patients in general.

In a direct comparison between different approaches to remotely assess symptoms in PwPD, video-based assessments were generally better accepted than sensor-based assessments and were described by patients as more easy to integrate into daily life^25^. Additionally, the vTUG can be completed with minimal time and effort. This indicates that the method could be employed not only for the purpose of conducting research, but also as a potential instrument for the measurement of evolving symptoms within the context of therapeutic intervention.

The TUG has a good test-retest reliability^30^, but inter-session reliability may reduce with increasing time (≥2 months) between longitudinal assessments^31^. Reliability can be further increased by averaging performance of three trials^32^. In the present study, assessments were performed 1x/week and each vTUG consisted of three repetitions. We did not observe significant time variations in those consecutively recorded repetitions, suggestiongs that they could be reduced to one repetion in PwPD. In some patients, we found time differences of up to 10 seconds from week to week, while others showed relatively stable times throughout the study. The simplicity of the vTUG task and app-supported recordings could allow for shorter intervals between recordings in future trials. This could help differentiate more reliably between psychometric weaknesses of the TUG and real changes in disease severity.

### Questionnaires

The mean SUS score for interfaces is 68.5 ^33^. Our score of 75,5 is therefore above average. Even with smaller sample sizes the SUS showed that it still can provide valid scores^34^, so we consider this finding meaningful, consistent with the low attrition rate observed in our cohort.

The UEQ offers the possibility to give some indication of the areas where improvements will have the greatest impact^35^. The UEQ exhibited an above-average performance in only a single category, with the remaining five categories demonstrating performance below the mean. This may initially appear to be an unfavourable outcome. Yet, it is important to note that the questionnaire was not designed exclusively for medical applications. Our objective was to create an app that is as user-friendly as possible for as many patients as possible. It is to be expected that this may result in a reduction in other valuation categories. For example, the weekly video recordings entail additional work without any direct benefit. This could explain the poor result in the stimulation category. However, the high rating in Perspicuity indicates that patients did not encounter difficulties in becoming familiar with the app and were able to learn how to use it with ease.

### Improvement Suggestions

The vTUG performance was influenced by various factors such as the chair, footwear, and location^18^. To ensure standardisation of the videos in future studies, it will be critical to make sure patients use the same set-up throughout all of them, and as mentioned specifically for chair, shoes, location and light. An example of an improvement could be the option to take a photo of their setup during their first session and store it in the app as a reminder. This would allow patients to remember which items were used during the initial attempt. Additionally, regular video checks should be conducted especially during the initial weeks to make sure patients are both performing and recording the vTUG correctly.

### Limitations

This study had several limitations related to the controlled inpatient setting. The majority of patients had a less severe form of PD, indicated by a Hoehn and Yahr stage of 1 or 2. The number of patients with a higher stage of PD is not representative, and it is likely that they have more difficulty navigating through the app and following the task without additional support of a spouse or carer.

Additionally, the study participants were primarily patients with prior technical experience. Those who did not feel comfortable using smartphones or lacked relatives who did so may have been unwilling to participate. As the study has no impact on the patients’ ongoing therapy, it did not increase the motivation of patients to try it even with small technical experience. While it could be concluded that this approach may only be suitable for a limited number of PD patients, this number is expected to grow rapidly in the future with the growing use of technical devices in aging people^36^, and as some of the positive feedback on ease of use as illustrated in Figure 7.

**Figure 7:**
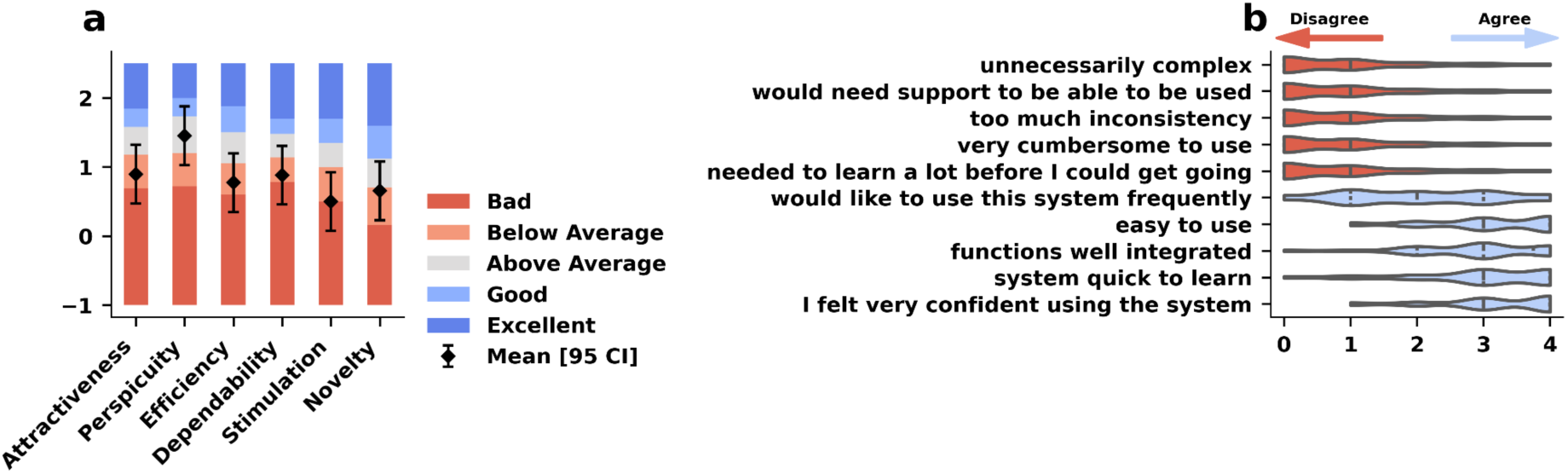
Usability testing revealed above-average perspicuity, emphasizing the ease of learning how to conduct the vTUG, but also indicated some room for improvement in user satisfaction and design. *a. Overall UEQ ratings for the vTUG (black diamonds and whiskers) across the six established domains compared against benchmarks (coloured bars). The measured scale means (± standard deviation) are presented in relation to existing values from a benchmark data set. b. Ratings for the 10 SUS items on a 5 item Likert scale (strongly disagree, disagree, neutral, agree, strongly agree)*.

To address these methodological weaknesses, future research should take into account the aforementioned limitations and ensure support for the digital literacy of all.

## Conclusion

The vTUG represents a promising approach to obtaining regular data on symptom severity and fluctuations in the home environment. It is a time-saving and simple method for patients to regularly transmit data to their doctor and has the potential to reduce visit burden for future clinical trials. However, when implementing this approach, it will be essential to pay attention to standardization with regard to factors such as the set-up and the time interval between the last medication intake.

## Data Availability

All data produced in the present work are contained in the manuscript.

## Notes

### Competing Interest Statement

MGE, CM, AMG, LCF, EHD are affiliated with Aparito Ltd, a wholly owned subsidiary of Eli Lilly and Company.

### Funding Statement

Aparito Ltd, a wholly owned subsidiary of Eli Lilly and Company.

### Author Declarations

The study was approved by the ethics committee of Technische University Dresden, Germany (BO-EK-149032021_3) and written informed consent was obtained from all patients before the participation.

